# Derivation of a clinical prediction rule for the early identification of patients at risk for developing post-COVID-19 condition when presenting to emergency departments with an acute SARS-CoV-2 infection

**DOI:** 10.64898/2026.01.06.26343467

**Authors:** Patrick M. Archambault, Rhonda J. Rosychuk, Martyne Audet, David Seonguk Yeom, Jeffrey P. Hau, Lorraine Graves, Simon Décary, Ivy Cheng, Jeffrey J. Perry, Steven C. Brooks, Laurie J. Morrison, Raoul Daoust, Hana Wiemer, Patrick T. Fok, Andrew D. McRae, Kavish Chandra, Michelle E. Kho, Bilkis Vissandjée, Matthew Menear, Eric Mercier, Samuel Vaillancourt, Dianne Zakaria, Phil Davis, Jean-Sébastien Paquette, Murdoch Leeies, Susie Goulding, Elyse Berger-Pelletier, Corinne M. Hohl, Canadian COVID-19 Emergency Department Rapid Response Network (CCEDRRN) investigators, Network of Canadian Emergency Researchers

## Abstract

**Background:** COVID-19 patients seen in an emergency department (ED) are at high risk of complications including post-COVID-19 condition (PCC), commonly known as Long COVID. As evidence is emerging concerning the efficacy of early post-acute rehabilitation and therapeutic interventions, early ED identification supported by a clinical prediction rule, combined with appropriate outreach and health education, could contribute to alleviating the burden of the disease on health systems and positively impact the quality of life of those living with the post-COVID-19 condition. This study aimed to derive and validate a clinical prediction rule to identify adult ED patients at high risk of developing PCC three months after an acute infection.

**Methods and findings:** This derivation and validation study used data from an observational cohort recruited from 33 hospitals in five Canadian provinces participating in the Canadian COVID-19 Emergency Department Rapid Response Network (CCEDRRN). We included adults (age ≥18 years) with confirmed COVID-19 who presented to the ED of a participating site between October 18, 2020, and October 11, 2022. We randomly assigned participants to derivation (75%) or validation (25%) datasets, and prespecified clinical variables as candidate predictors. We used a fast step-down logistic regression to reduce the model to key predictors for our clinical prediction rule. Validation was planned only if the derived rule had an AUC of at least 80% to support clinically useful discrimination characteristics to separate those who will develop PCC from those who will not. Of 6,070 eligible patients, 2,511 (41.4%) reported PCC symptoms at three months. Our derived clinical prediction rule included nine risk factors (female sex, higher arrival respiratory rate, comorbidities (rheumatologic disorder and mental health condition), acute symptoms (sputum production, dizziness, diarrhea, chest pain, and fatigue)) and one protective factor (self-reported South Asian race). In derivation, the optimism-corrected area under the curve was 0.626 (95% confidence interval [CI] 0.610–0.643). Age and vaccination status were not retained in the final clinical prediction rule. The rule was only slightly better than chance and deemed not accurate enough to meaningfully guide decision-making in the ED. Therefore, we did not proceed to examine its performance in the validation cohort.

**Conclusions:** Despite rigorous methodology, we were unable to derive a clinical prediction rule with sufficient accuracy to predict PCC in emergency department patients at the time of the acute infection. However, we did identify several factors associated with the development of PCC that can guide future studies about the causes of PCC. The ambiguous nature of the current PCC diagnostic criteria and the extended follow-up pose challenges for deriving a useful clinical decision rule. Further research integrating comprehensive surveillance systems and biomarker data may also enhance prediction accuracy and refine personalized management strategies in the emergency department setting.

## Introduction

Emergency departments around the world continue to play a critical role in evaluating and managing COVID-19 infections.(1,2) Between April 2022 and March 2023, Canadian EDs saw >220K visits for COVID-19.(3) The majority of people (69%) who went to the ED were discharged home, while 26% were admitted to hospital.(4) Now that community-based testing is less available,(5) more people with upper respiratory infections are presenting to EDs seeking diagnosis and treatment.(6,7) EDs also see the sickest COVID-19 patients who are at the highest risk of complications including post-COVID-19 condition (PCC), also commonly known as Long COVID.(8) The World Health Organization (WHO) defined PCC in October 2021 as a condition that “*occurs in individuals with a history of probable or confirmed Severe Acute Respiratory Syndrome Coronavirus-2 (SARS-CoV-2) infection, usually 3 months from the onset of COVID-19 with symptoms that last for at least 2 months, that cannot be explained by an alternative diagnosis*”.(9–12) While common symptoms of PCC can include fatigue, shortness of breath and cognitive dysfunction, over 200 different symptoms have been reported that can have an impact on everyday functioning. Based on recent prevalence estimates, more than 400 million individuals could be living with PCC worldwide.(13,14) People with PCC have increased primary care visits, hospital admissions and mortality in the months following infection.(15,16) ED patients are at higher risk of PCC compared to patients with COVID-19 who don’t present to the ED, with 40% of ED patients with a SARS-CoV-2 infection reporting PCC at three months compared to 15% of patients in the community.(8,17) For patients and ED clinicians alike, recognizing which ED patients are at high risk of developing PCC at the time of the acute infection may be important for early follow-up, and potential preventative and therapeutic interventions. As evidence is emerging concerning the efficacy of early post-acute rehabilitation(18–23) and potential therapeutic interventions such as metformin(24), probiotics(25) and antivirals(26), early ED identification, combined with appropriate outreach and patient education, could contribute to alleviating the burden of the disease on health systems and positively impact patients’ quality of life.

Multiple risk factors for developing PCC have been reported, including age, sex, premorbid risk factors,(27) hospitalization, vaccination status,(28) circulating variant, reinfection,(29) and prolonged intensive care stays.(30) Existing prediction models have been developed using retrospective administrative data and machine learning approaches(31) or have been developed using data from unvaccinated pre-Omicron patients.(32) For these reasons, there remains a need to develop a clinical prediction rule for use in the ED using readily available data to support decision-making about referral for early preventative and therapeutic interventions.(33,34) This study aimed to derive and validate a clinical prediction rule (CPR) to identify adult ED patients at high-risk of developing PCC three months after an acute infection.

## Methods

### Design and setting

This observational multicenter cohort study recruited patients from 33 EDs across five Canadian provinces (British Columbia, Saskatchewan, Ontario, Québec and Nova Scotia) that participated in the Canadian COVID-19 Emergency Department Rapid Response Network (CCEDRRN, S1 Table).(35) We recruited consecutive consenting patients who visited a participating ED between October 18, 2020, and October 11, 2022. The University of British Columbia Clinical Research Ethics Board (H20-01015) as well as the research ethics boards of all participating institutions approved this study with a waiver of informed consent for enrollment and collection of retrospective data from hospital charts, with permission to contact patients to obtain their consent for follow-up phone interviews in French or English. Model development and reporting followed Transparent Reporting of a Multivariable Prediction Model for Individual Prognosis or Diagnosis (TRIPOD) standards (S2 Table).(36) Our protocol is registered with clinicaltrials.gov (<u>NCT04702945</u>).

CCEDRRN’s Patient Engagement Committee consisted of 11 members from 6 provinces across Canada (BC, AB, SK, MN, ON, QC, NS). The committee was composed of 6 women and 5 men. Members had lived experience with COVID-19 and PCC and were engaged from this study’s inception onwards. They participated in generating the original research question, grant writing, edited consent and data collection forms to ensure readability and acceptability in both French and English and across diverse social, cultural, and ethnic contexts, assisted with developing and refining study questions, interpreted study findings, and reviewed and edited the manuscript. We report our patient engagement strategy(37) using the GRIPP2-SF guideline (S3 Table).(38)

### Participants

We included consecutive consenting participants ≥18 years of age who tested positive for SARS-CoV-2 infection, defined as ≥1 positive SARS-CoV-2 nucleic acid amplification or rapid antigen tests in the 14 days before or during the ED visit, or in the 14 days after hospital admission. Participants had to be able to communicate in French or English, and complete at least one Post-COVID Condition Assessment Questionnaire over the phone (PCCAQ; S1a and S1b Files). We excluded patients who died, were hospitalized, or were out of the country at the time of follow-up, who could not be contacted after 5 attempts, were unable to communicate due to language or cognitive barriers, who did not complete the interview, or provided incomplete vaccine information.

### Data Collection

Trained research assistants: (i) abstracted data on SARS-CoV-2 tested patients including their baseline comorbidities by chart review,(35) (ii) attempted to contact patients up to 5 times to obtain verbal consent for phone follow-up at 6 months following the ED visit, (iii) collected sociocultural and demographic variables including age, sex, gender, race and ethnicity, as well as baseline level of fitness, and self-reported SARS-CoV-2 vaccination status,(39,40) (iv) documented any self-reported new or repeat SARS-CoV-2 infections, and (v) documented ongoing or resolved symptoms consistent with PCC using the PCCAQ. Research assistants were trained to only document new symptoms that developed since the ED index visit. We developed the PCCAQ based on the WHO PCC case definition and case report form(41) in collaboration with patient partners, PCC experts, emergency physicians, rehabilitation specialists, and public health policy makers. We piloted a first version of the PCCAQ (PCCAQ version 1.0, S1a File) in English and French with CCEDRRN patient partners and the first 100 participants. Then, we used this version of the PCCAQ with all patients whose index visit occurred between October 18, 2020, and February 28, 2022. For all patients with index visits after February 28, 2022, we used a new and simplified version of the PCCAQ that was improved to help patients determine the duration of their symptoms using open-ended questions instead of asking them to categorize the duration of their symptoms using predetermined multiple-choice options (PCCAQ version 2.0, S1b File). This new PCCAQ version helped reduce the time to complete the questionnaire. Phone follow-ups for this study occurred between November 16, 2021, and August 31, 2023.

### Outcome

The primary outcome was the presence of at least one self-reported symptom consistent with PCC at three months after their index ED visit. We defined PCC according to the WHO definition(12) as patients reporting new onset symptoms not attributable to another diagnosis that started within three months of their index ED visit and lasted at least two months. Symptoms had to remain present at or past the three-month mark. The list of compatible symptoms was based on the symptoms found in the *Global COVID-19 Clinical Platform Case Report Form (CRF) for Post COVID condition (Post COVID-19 CRF).*(41)

### Data Analysis

We chose candidate predictor variables based on literature review, clinical acumen, patient partner lived/living experience, and previously published results on predictors of reporting symptoms consistent with PCC using CCEDRRN data.(9) In all, 21 candidate predictor variables were selected (S4 Table), including age, sex, pre-existing health conditions (active/past malignant neoplasm, asthma, chronic lung disease (e.g., COPD), lung fibrosis, atrial fibrillation, chronic kidney disease, chronic lung disease, chronic neurological disorder (e.g., stroke, seizure disorder), congestive heart failure, coronary heart disease, diabetes, dialysis, dyslipidemia, hypertension, hypothyroidism, mild liver disease, moderate/severe liver disease, obesity, organ transplant, psychiatric condition, rheumatological disorder), race, education, acute symptoms and severity at index ED visit, vaccination status, and substance misuse.

### Model development

We randomly assigned patients to derivation or validation datasets, with the goal of assigning 75% of eligible patients and primary outcome events to the derivation dataset, and the remaining to the validation dataset. Summary statistics were calculated for the candidate predictor variables which were examined for clinical significance and availability at the time of the ED visit. Lab and imaging tests were excluded because they delay triage and decision-making, potentially contribute to overcrowding and are not accessible in resource-limited settings.(42,43) We used nine multiple imputations for variables with missing data if data could not be reasonably assumed (S4 Table): arrival heart rate, arrival respiratory rate, arrival blood pressure, arrival temperature, Canadian Triage and Acuity Scale). Missing data for self-declared race and education level were not imputed because their missingness was not at random.

We assessed collinearity among variables using clustered Spearman correlations and variance inflation factors, and no significant issues were identified in either analysis. Additionally, we evaluated redundancy among variables with a cut-off R^2^ of 0.75, and no variables were excluded based on this criterion. In the initial logistic regression model, we included all selected candidate predictor variables with restricted cubic spline functions with three knots fit for continuous variables. We assessed the strength of associations using an ANOVA plot to allocate the degrees of freedom to each variable in the model. We also considered interactions between sex and other candidate variables. We used a fast step-down procedure to reduce the model to key predictors.

We used internal bootstrap validation with 1,000 bootstrap samples to assess for model overfitting and calculate an optimism-corrected area under the receiver-operating characteristic (ROC) curve (AUC) (S5 Table). We planned to use the final reduced model to create an easy-to-use points score and then calculate the sensitivity and specificity at different point thresholds, along with discrimination and calibration of the score. Additionally, validation of our clinical prediction rule was planned only if the derived rule had an AUC of at least 80% and acceptable performance characteristics.(44) We conducted all analyses in R using the rms package.(45–47) To ensure patient privacy, a cell size restriction policy prohibited reporting counts of <5.

### Sample size

Assuming an event rate of 34% to 39% based on previous data,(8) shrinkage of 0.9, and a conservative Cox-Snell R-squared of 0.3, about 9.6 events per degree of freedom were required for reliable prediction modeling in the derivation cohort.(48) We selected 21 candidate predictors based on clinical experience and previous literature.(8) The 21 candidate predictor variables had 72 degrees of freedom, which indicated that 692 events were required. In the derivation cohort, there were 4,566 patients with 1,879 (41.2%) who experienced PCC representing 1,879 events, which exceeded the number of events required.

## Results

We assessed 23,348 patients with COVID-19 who visited a participating ED between October 18, 2020, and October 11, 2022 (Fig 1).

**Figure 1.**
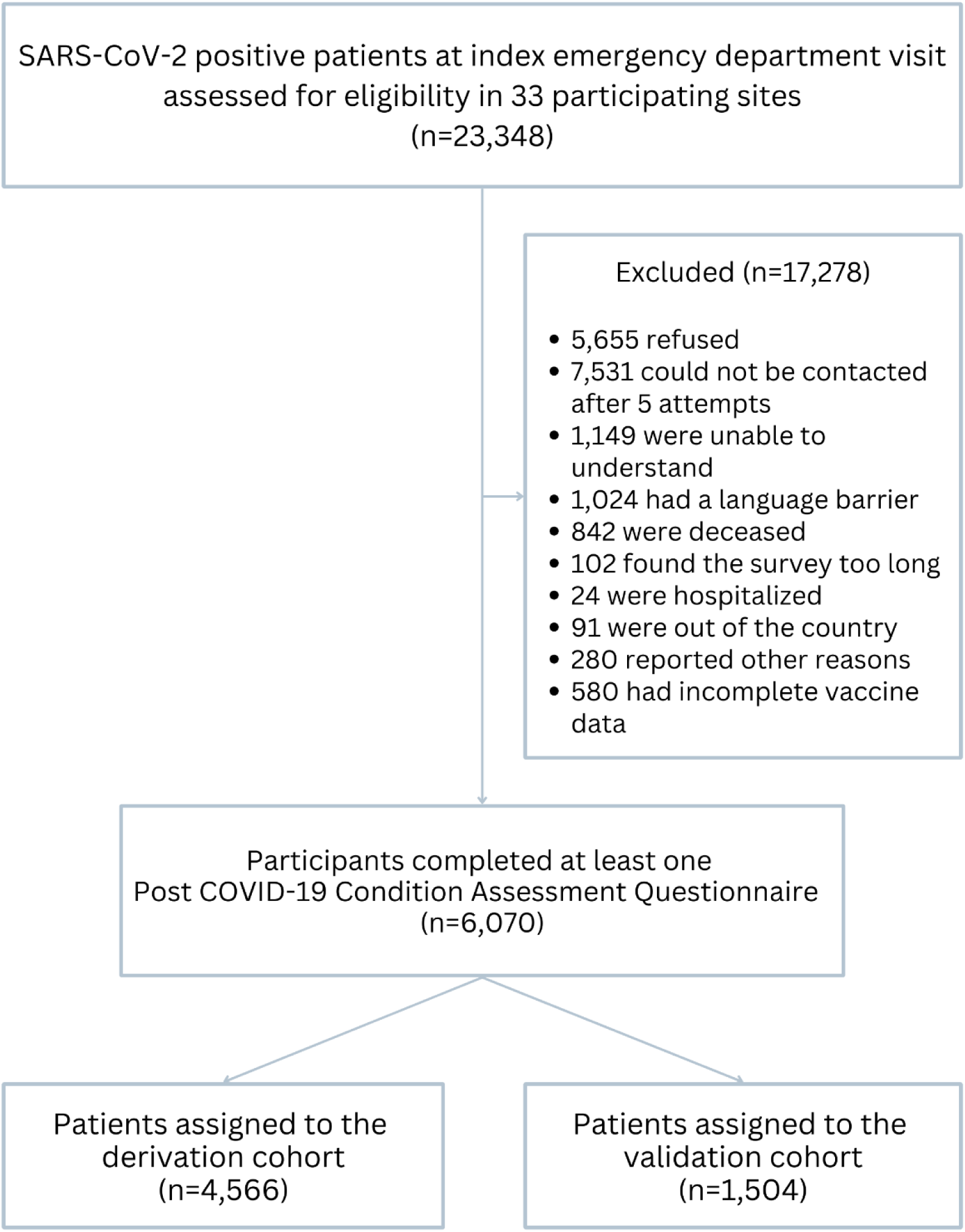
Flow diagram showing included and excluded emergency department patients in the derivation and the validation cohorts.

We excluded 17,278 patients who met at least one exclusion criteria and included 6,070 patients in our analyses. Of 6,070 patients, 2,511 (41.4%) reported symptoms consistent with PCC at three months after their index ED visit date. The characteristics and outcomes of patients in the derivation (n = 4,566) and validation (n = 1,504) cohorts are shown in Table 1. The derivation and validation cohorts had similar baseline characteristics. The three-month prevalence of PCC in both cohorts was similar (41.2% vs. 42%).

**Table 1.** Baseline patient characteristics and outcomes of SARS-CoV-2 positive patients presenting to emergency departments in the derivation and validation cohorts (N=6,070).

|  | **Derivation**<br>**(n = 4566)** | **Validation**<br>**(n = 1504)** |
| --- | --- | --- |
| **Age in years, Mean (SD)** | 51.1 (17.7) | 51.1 (17.9) |
| **Female, n (%)** | 2387 (52.3) | 773 (51.4) |
| **Province, n (%)** |  |  |
| British Columbia | 2197 (48.1) | 740 (49.2) |
| Québec | 1966 (43.1) | 625 (41.6) |
| Ontario | 308 (6.7) | 101 (6.7) |
| Saskatchewan | 33 (0.7) | 20 (1.3) |
| Nova Scotia | 62 (1.4) | 18 (1.2) |
| **Arrival mode, n (%)** |  |  |
| Self | 3141 (68.8) | 1006 (66.9) |
| Ambulance or police | 1425 (31.2) | 498 (33.1) |
| **Arrival heart rate, beats /min, mean (SD)** | 94.7 (18.2) | 94.4 (19.0) |
| **Arrival systolic blood pressure, mmHg, mean (SD)** | 130.7 (20.8) | 130.9 (20.8) |
| **Arrival respiratory rate/min, mean (SD)** | 20.2 (5.4) | 20.0 (5.2) |
| **Arrival temperature in degrees Celsius, mean (SD)** | 37.2 (0.9) | 37.2 (0.8) |
| **Presence of respiratory distress, n (%)** | 634 (13.9) | 221 (14.7) |
| **COVID-19 acute symptoms, n (%)** |  |  |
| Cough | 2831 (62.0) | 917 (61.0) |
| Shortness of breath (dyspnea) | 2262 (49.5) | 740 (49.2) |
| Fever | 2038 (44.6) | 685 (45.5) |
| Chest pain (includes discomfort or tightness) | 1705 (37.3) | 578 (38.4) |
| Fatigue/malaise | 1563 (34.2) | 527 (35.0) |
| **Most common comorbid conditions, n (%)** |  |  |
| Hypertension | 1141 (25.0) | 392 (26.1) |
| Dyslipidemia | 708 (15.5) | 260 (17.3) |
| Diabetes | 694 (15.2) | 215 (14.3) |
| Asthma | 475 (10.4) | 144 (9.6) |
| Psychiatric Condition/Mental Health Diagnosis | 470 (10.3) | 147 (9.8) |
| **Oxygen required in Emergency Department, n (%)** | 723 (15.8) | 240 (16.0) |
| **Vaccine status, n (%)** |  |  |
| 0 or 1 dose | 2392 (52.4) | 756 (50.3) |
| ≥ 2 | 2174 (47.6) | 748 (49.7) |
| **Emergency Department disposition, n (%)** |  |  |
| Discharged | 3232 (70.8) | 1063 (70.7) |
| Admitted | 1318 (28.9) | 438 (29.1) |
| Left against medical advice | 9 (0.2) | < 5 |
| Other | 7 (0.2) | < 5 |
| **Pandemic Wave, n (%)** |  |  |
| Prior to Omicron (before November 28, 2021) | 2336 (51.2) | 739 (49.1) |
| During Omicron (on or after November 28, 2021) | 2230 (48.8) | 765 (50.9) |
| **Post-COVID-19 Condition prevalence, n (%)** |  |  |
| 3 months | 1879 (41.2) | 632 (42.0) |

SARS-CoV-2: Severe Acute Respiratory Syndrome Coronavirus-2; SD: Standard deviation.

In the derivation cohort, the step-down procedure produced a final reduced model with ten variables (Table 2).

**Table 2.** Adjusted associations between predictor variables and post-COVID-19 condition three months after emergency department index visit.

| Variable | Estimate (SE) | OR (95% CI) |
| --- | --- | --- |
| **Female** | 0.40 (0.06) | 1.49 (1.32, 1.68) |
| **Arrival respiratory rate**, breaths/min<br>(30 breaths/min vs 18 breaths/min)* | 0.43 (0.04) | 1.54 (1.28, 1.85) |
| **COVID-19 related acute symptoms** |  |  |
| Sputum production | 0.39 (0.10) | 1.48 (1.22, 1.78) |
| Dizziness/Vertigo | 0.33 (0.10) | 1.39 (1.14, 1.71) |
| Diarrhea | 0.28 (0.06) | 1.32 (1.12, 1.56) |
| Chest pain (includes discomfort or tightness) | 0.27 (0.06) | 1.31 (1.16, 1.49) |
| Fatigue/malaise | 0.21 (0.07) | 1.23 (1.08, 1.40) |
| **Comorbidities** |  |  |
| Rheumatologic disorder | 0.43 (0.13) | 1.54 (1.20, 1.97) |
| Psychiatric Condition/Mental Health Diagnosis | 0.35 (0.10) | 1.41 (1.16, 1.72) |
| **South Asian** | -0.60 (0.10) | 0.55 (0.45, 0.67) |

SE: Standard error; OR: Odds ratio; 95% CI: 95% Confidence interval; \* These respiratory rates represent single respiratory rate values selected to predict the post-COVID-19 condition (PCC) based on the splined respiratory rate. They show that higher arrival respiratory rates (i.e., 30 vs. 18) are associated with an increased risk of PCC (S1 Fig.).

Age, vaccination status and education were not retained in the final model because they were not significantly associated with PCC in our multivariable model (S4 Table). Internal bootstrap validation demonstrated no serious overfitting issue with the final variables. We note that age was an important factor in our internal bootstrap validation sample but was not included in our final model using the modeling process (S5 Table). The derived model had an optimism-corrected AUC (95% CI) of 62.6% (61.0%-64.3%) (Fig 2). Interaction effects between candidate predictors and age did not significantly improve the final model.

**Figure 2.**
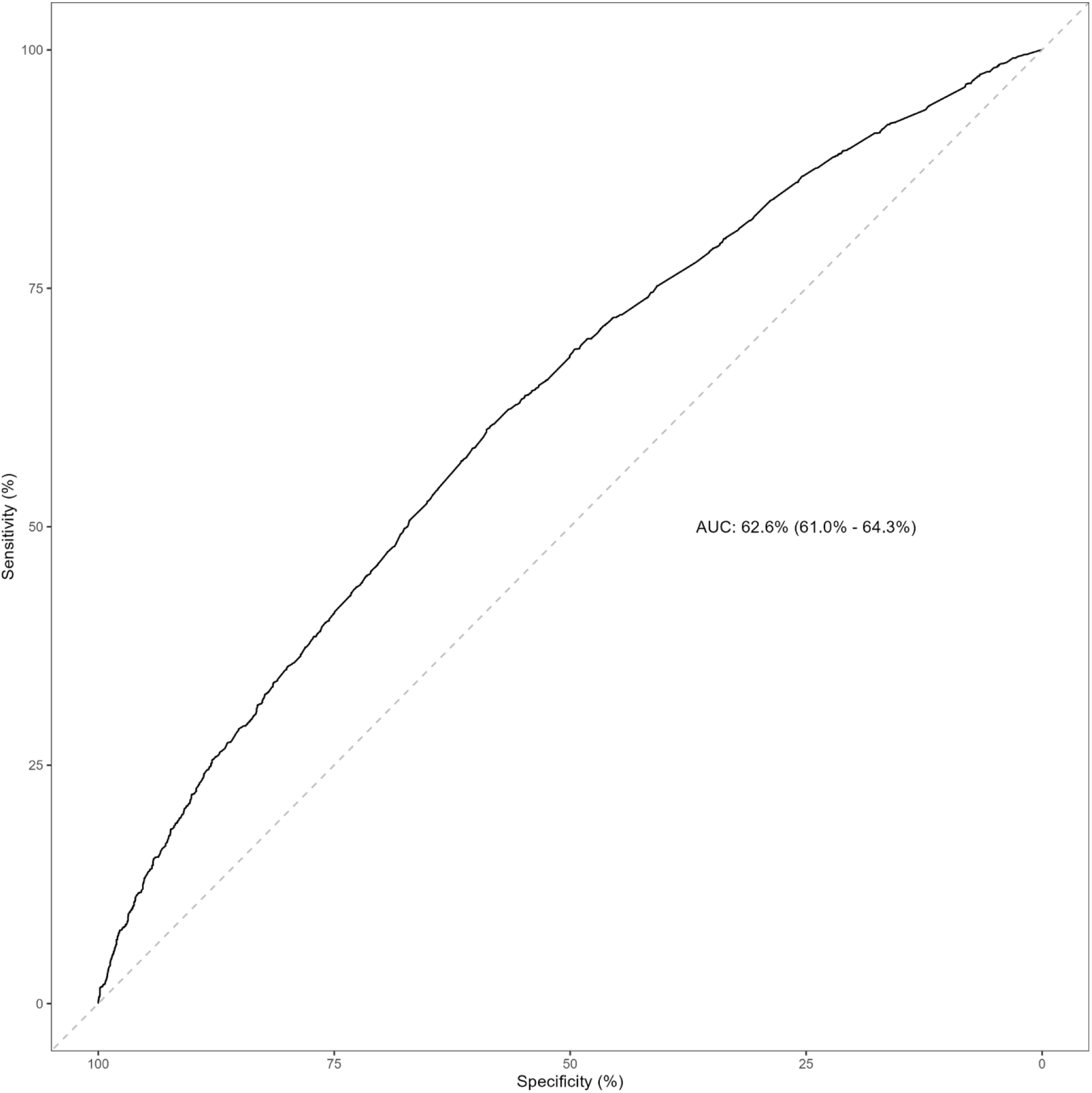
Area under curve (AUC) of the derived model.

The rule was only slightly better than chance and deemed not accurate enough to meaningfully guide decision-making in the ED. Therefore, we did not proceed to examine its performance in the validation cohort.

## Discussion

Our goal was to develop a simple parsimonious prediction rule that could be used at the bedside to predict the development of PCC in adult ED patients. Despite the use of state-of-the-art clinical prediction rule methodology, we were unable to derive a clinical prediction rule that had acceptable performance characteristics to move forward to validation. We identified several risk factors that increased the risk of developing PCC including female sex, respiratory rate, previous rheumatologic disorder or mental health diagnosis, and reporting sputum production, dizziness/vertigo, diarrhea, chest pain or fatigue at the time of the acute infection, but none were strong predictors like those identified in other COVID-19 models.(8,49) The only protective factor was self-reported South Asian race. Age and vaccination status were not retained as risk factors in our final model.

Even though we were not able to derive the intended clinical prediction rule, our study has several strengths. First, only a few studies systematically followed consecutive SARS-CoV-2 tested patients and integrated clinical data from the acute infection with patient-reported information.(50–52) Second, we rigorously applied the WHO definition using specific time cut-off points and asked patients to discern new versus chronic symptoms, improving the specificity of the patients identified as having PCC. Third, our inclusion of patients from 33 participating centers in Canada from five provinces ensured external validity across different care settings and representativeness of the most vulnerable patients seen in the Canadian healthcare system. Fourth, our data spans both pre- and post-Omicron pandemic periods increasing its applicability to multiple circulating variants.(53) Fifth, this study was developed with the participation of patient partners who provided guidance in its development, its conduct and interpretation.

Compared to existing published PCC prediction rules,(31,32,54–58) our results are similar with poor to fair validation performance statistics with AUCs reported between 0.62 and 0.79.(44) One model developed in the United States(59) reports an AUC of 0.87, but the PCC definition used is unclear and the model was developed with machine learning built to optimize the AUC using data from a curated administrative database decreasing its applicability in clinical practice. Another model developed using data from ED patients in Mexico produced an AUC of 0.70 based on a symptom severity score,(32) but it was derived without internal or external validation using data from unvaccinated pre-Omicron patients, therefore limiting its usefulness with current post-Omicron ED patients. A study from China derived and validated a clinical prediction rule to predict ongoing symptoms at 4-8 weeks post infection for hospitalized COVID-19 patients.(31) Their rule based on age, diabetes, chronic kidney disease, vaccination status, and inflammation biomarker tests (procalcitonin, leukocytes, lymphocytes, interleukin-6, D-dimer) had fair discrimination in both derivation (AUC: 0.762) and validation (AUC: 0.713). However, this rule was not developed for use at the bedside in the ED at the time of acute infection when preventative treatments may be most effective.(31) In the context of resource and time limited ED care, a rule that relies on multiple time-consuming laboratory results would be impractical, and costly.(60)

Creating an effective PCC clinical prediction rule for ED patients is difficult, as evidenced by the many attempts that have fallen short of the desired performance levels. The WHO definition for PCC is very broad and remains hard to operationalize for the development of a clinical prediction rule.(11) The current WHO definition allows for misclassification of the outcome to include many non PCC patients (e.g., other new onset respiratory or neurological illnesses) in addition to true PCC patients. The inherent uncertainty about the definition of PCC leads to ambiguity in defining the outcome for prediction modeling and possibly a low AUC value. The definition is particularly difficult to apply in the case of relapsing symptoms, deterioration of preexisting undiagnosed conditions, undocumented COVID-19 infections and reinfections, multisystem presentations, and currently includes a myriad of non-specific symptoms. Although our questionnaire was built to detect any new symptoms since the ED index visit, PCC remains a clinical diagnosis that relies on the exclusion of all other causes. As our study demonstrates, ruling-in PCC remains a challenge because the diagnostic criteria are not specific, and it remains difficult to differentiate new symptoms related to PCC from those of other new conditions that can be diagnosed concomitantly.

Nonetheless, the development of a clinical prediction rule that could support the identification of high-risk patents for PCC, and to initiate treatment or facilitate referrals for early preventative and therapeutic interventions is still very relevant. The findings of our study reveal several challenges and insights about the development of a clinical prediction rule for PCC in the ED setting that will need to be addressed in the future. The WHO definition for PCC, while comprehensive, relies on the presence of non-specific symptoms that overlap with many other conditions (e.g., new onset COPD) that make a definitive diagnosis of PCC difficult to confirm and prevents the derivation of a highly performing prediction rule. The National Academies of Sciences, Engineering, and Medicine has recently proposed a new unified definition for PCC.(61) Although several improvements are proposed with inclusive parameters regarding timing of symptoms, inclusion of asymptomatic and unproven COVID infections, and the integration of important patient-oriented features, the current absence of specific biomarkers adds to the reliance on interpretation of non-specific symptoms for a clinical diagnosis that will make the development of a prediction rule for PCC based on this new definition even more difficult.

Although we did not succeed in deriving a multivariable prediction rule with adequate performance characteristics, our study identified individual risk factors similar to ones found in other studies and highlighted new ones that merit further investigation.

First, our results concur with multiple systematic reviews that found that female sex is a risk factor for PCC.(62–66) Potential explanations include the role of sex hormones (67), higher innate immune responses in females,(68) and social factors and gender biases making it more acceptable for women to disclose pain and distress compared to men.(69–71)

We also found that premorbid rheumatologic disorders were also an independent risk factor for PCC. Many authors have found that COVID-19 is linked with hyperinflammation and antibody production that can lead to rheumatologic disorders.(72,73) However, few authors(74–76) have been able to clearly demonstrate that patients with rheumatologic disorders are at higher risk of PCC because of overlapping symptoms and pre-morbid immunosuppression potentially decreasing production of proinflammatory cytokine production.

In line with previous systematic reviews, we also found that preexisting mental health problems increased the risk of PCC.(62,64,66) An international cohort study of nurses and their offspring found premorbid psychological distress, including symptoms of depression, symptoms of anxiety, fear about COVID-19, loneliness, and perceived stress, was strongly associated with risk of PCC.(77) Psychological distress at the time of viral infections has been associated with longer and more severe upper respiratory tract infections.(78,79) Distress is also associated with chronic systemic proinflammatory cytokines.(80–82) Although these cytokines have been proposed as possible causes of neurological, psychiatric and neurocognitive PCC symptoms, further research into the pathophysiology of PCC and the role of mental disorders is needed.(83,84)

Consistent with another Canadian study that used machine learning to derive a predictive model to identify PCC patients,(85) our study highlighted an association between three acute COVID-19 symptoms (sputum production, chest pain, and fatigue) and the development of PCC at three months. Aside from studies that showed that the total number of acute COVID-19 symptoms were predictive of PCC, none of the existing PCC prediction models specifically included acute-phase diarrhea and/or vertigo as predictors for PCC.(54–56,86) A meta-analysis about the gastrointestinal manifestations of COVID-19 reported that 12.5% of patients with COVID-19 had diarrhea.(87) In this meta-analysis, patients with acute diarrhea had higher stool SARS-CoV-2 RNA positivity and viral load than those without diarrhea. Prolonged shedding of stool viral RNA which lasted up to a month after the acute illness and after the end of respiratory viral RNA shedding was also observed in 70% of patients. Moreover, dysbiosis (changes in the gut microbiome and gut wall integrity) has been linked to the pathophysiology of PCC by increasing systemic inflammation.(88–90). Acute diarrhea may thus be associated with both dysbiosis and increased risk of PCC.(91) As for vertigo and dizziness, another study reported that one sixth of patients with COVID-19 presented with vertigo or dizziness.(92) While neurological symptoms during the acute phase such as anosmia have been associated with an increased risk of PCC,(8) acute vertigo or dizziness have not been consistently identified as independent predictors.(93,94)

Higher arrival ED respiratory rate was associated with increased risk of PCC in our study. This aligns with prior studies that have shown that more severely affected COVID-19 patients requiring hospital admission or intensive care were at higher risk for PCC.(8,60,95–97) This is also consistent with our results that found that presenting acute respiratory symptoms (sputum production and chest pain) increased the risk of PCC.

In agreement with other studies,(98) we found a protective effect of being South Asian. A systematic review of 102 studies about PCC dyspnea pooling results from 42,872 patients around the world found that studies from Asia seem to report less post COVID-19 dyspnea(99) Many explanations have been proposed including the predominance of vegan and antioxidant-rich diets favored by South Asians that may have anti-inflammatory properties,(100,101) lower social acceptability in reporting mental health symptoms(102) language barriers,(103) and higher vaccination rates in Canadian South Asian population.(104)

Contrary to other COVID-19 outcomes (e.g., hospitalization, respiratory failure and mortality(49,105)), we did not find an association between age and PCC in our derivation cohort. As described in many reviews,(100,106) age is inconsistently reported as a risk factor across studies and is most likely confounded with age associated comorbidities.(107) Other studies have shown that middle-aged women are at higher risk of PCC, so increasing age might have a protective effect or bimodal effect.(108) Another potential explanation could be selection bias due to more older adults lost to follow-up, unable to consent or deceased.(109,110)

As we have identified in previous works,(8) we did not find a protective effect for vaccination status on PCC. Although the protective effect of SARS-CoV-2 vaccination on the development of PCC is largely recognized,(108,111–113) there are several factors that may explain our results. The COVID-19 pandemic was characterised by strong disruptions of health care, confusing information, lack of access to reliable vaccination data,(114) different COVID-19 variants of concern, lockdowns, and variations in timing of vaccine programs across Canadian provinces, all of which will have influenced the consolidated risk to develop PCC for which we could not control in our analyses. As the most effective way to prevent PCC is to prevent SARS-CoV-2 infection (e.g., vaccination, masking, social distancing, hand washing), it is likely that any measure taken to decrease the incidence of acute SARS-CoV-2 infection will in turn prevent the development of PCC.

Our study has several limitations. First, as previously stated, the WHO PCC definition is very broad and remains hard to operationalize for the development of a clinical prediction rule.(11) Although our questionnaire was designed to capture any new symptoms since the ED index visit, PCC remains a clinical diagnosis that relies on the exclusion of all other causes. Ruling-in PCC remains a challenge because the diagnostic criteria are not specific, and it remains difficult to differentiate new symptoms related to PCC from those of other new conditions that can be diagnosed concomitantly. Second, our PCC questionnaire was implemented without formal psychometric evaluation early during the pandemic when there was an urgency to capture PCC outcomes without any existing validated questionnaire. It was, however, co-developed with patient partners, experts in PCC and rehabilitation, then pilot-tested in French and English with a subset of patients, and implemented with training material to standardize its use. Because the outcome was measured based on self-reported symptoms collected via telephone follow-up, we lacked a gold standard to ensure that a patient was experiencing a relevant symptom. This may have introduced a response bias. Although our study is representative of the patients seen in Canadian EDs, following up with certain hard to contact populations such as homeless people, those living with cognitive disorders or those with language barriers remained a challenge. Third, we had to exclude cases of reinfection, which is now a recognized risk factor for PCC,(115) because our follow-ups were planned at designated time points. Despite rigorous methodology, we were unable to derive a clinical decision rule with sufficient accuracy to predict PCC in emergency department patients at the time of the acute infection. However, we did identify several factors associated with the development of PCC that can guide future studies about the causes of PCC. Future research should also focus on developing a more specific PCC definition and finding a biomarker that could support the definitive diagnosis of PCC. The ambiguous nature of the current PCC criteria and the long follow-up pose challenges for deriving a useful clinical decision rule. Further research, integrating more comprehensive surveillance systems and a more specific definition, may also enhance prediction accuracy and refine patient management strategies in the ED setting. The development of an accurate and user-friendly clinical decision rule still has the potential to significantly impact COVID-19 patient care and outcomes and could potentially help develop clinical prediction rules for other post-infectious conditions.

## Data Availability

Data is available on reasonable request. For investigators who wish to access CCEDRRN data, proposals may be submitted to the network for review and approval by the network's peer-review publication committee, the data access and management committee and the executive committee, as per the network's governance. Information regarding submitting proposals and accessing data may be found on the CCEDRRN website.

https://www.ccedrrn.com/

## Acknowledgments

We gratefully acknowledge the assistance of Amber Cragg and would like to thank The University of British Columbia clinical coordinating center staff, legal, ethics, privacy, and contract staff, and the research staff at each of the participating institutions in the network outlined in the S1 Table and S2 File. We would also like to thank CCEDRRN national coordinator Vi Ho, and provincial coordinators Josie Kanu (BC), Aimee Goss (SK), Connie Taylor and Vlad Latiu (ON), Corinne DeMone (NS), and Chantal Lanthier, Alexandra Nadeau, Xiaoqing Xue, and David Ianuzzi (QC) for their support in collecting data for this study. The network would not exist today without the dedication of these professionals. We would also like to thank Catherine Truchon and Marie-Claude Breton at the Institut national d’excellence en santé et en services sociaux for their collaboration with this project. Thank you to all the patient partners who shared their lived experiences and perspectives to ensure that the knowledge we cocreate addresses the concerns of patients and the public. We would like to thank Colleen McGavin for having supported the creation of our patient engagement committee at the inception of our network. Creating the largest network of collaboration across Canadian emergency departments would not have been possible without the tireless efforts of emergency department chiefs, and research assistants at participating sites. Finally, our most humble and sincere gratitude to all our colleagues in medicine, nursing, and the allied health professions who have been on the frontlines of this pandemic from day 1 staffing our ambulances, emergency departments, intensive care units, and hospitals bravely facing the risks of COVID-19 to look after our fellow citizens and after each other. We dedicate this network to you. The authors wish to acknowledge Long COVID Web for its role in supporting the completion of this Research Project. The authors no competing interests.

---

**S1 Table.** Patient enrollment in CCEDRRN’s registry across participating sites.

| Site Name | Province | Start Date* | End Date* | Number of consenting participants |
| --- | --- | --- | --- | --- |
| Vancouver General Hospital | British Columbia | 2021-01-29 | 2022-07-28 | 478 |
| Lions Gate Hospital | British Columbia | 2020-12-31 | 2022-07-31 | 259 |
| Saint Paul's Hospital | British Columbia | 2021-01-14 | 2022-07-31 | 384 |
| Mount Saint Joseph's Hospital | British Columbia | 2020-01-13 | 2022-07-31 | 197 |
| Surrey Memorial Hospital | British Columbia | 2020-12-08 | 2021-07-31 | 1196 |
| Royal Columbian Hospital | British Columbia | 2020-12-15 | 2021-05-30 | 146 |
| Abbotsford Regional Hospital | British Columbia | 2020-12-17 | 2021-05-31 | 166 |
| Eagle Ridge Hospital | British Columbia | 2020-12-15 | 2021-05-30 | 68 |
| Royal Inland Hospital | British Columbia | 2021-02-04 | 2021-05-24 | 17 |
| Kelowna General Hospital | British Columbia | 2021-01-04 | 2021-05-25 | 26 |
| St. Paul's Hospital | Saskatchewan | 2021-04-25 | 2021-12-25 | 15 |
| Royal University Hospital | Saskatchewan | 2021-04-11 | 2022-01-03 | 38 |
| Sunnybrook Health Sciences Center | Ontario | 2020-12-16 | 2022-02-23 | 118 |
| Ottawa Hospital – Civic Campus | Ontario | 2021-03-07 | 2021-05-21 | 26 |
| Ottawa Hospital – General Campus | Ontario | 2021-02-07 | 2021-05-21 | 24 |
| Kingston General Hospital | Ontario | 2021-01-04 | 2022-05-27 | 95 |
| Health Sciences North | Ontario | 2021-01-13 | 2022-07-30 | 81 |
| LHSC University Hospital | Ontario | 2021-01-24 | 2021-05-23 | 36 |
| Hotel Dieu Hospital | Ontario | 2021-04-16 | 2022-05-29 | 29 |
| Hotel-Dieu de Levis | Quebec | 2020-10-18 | 2022-07-20 | 446 |
| Jewish General Hospital | Quebec | 2020-11-29 | 2022-06-30 | 920 |
| Centre Hospitalier de l'Université Laval | Quebec | 2020-12-23 | 2021-04-10 | 11 |
| Royal Victoria Hospital | Quebec | 2020-12-01 | 2021-12-31 | 404 |
| Hôpital de l'Enfant-Jésus | Quebec | 2020-12-30 | 2022-01-31 | 324 |
| Hôpital du Saint-Sacrement, CHU de Québec | Quebec | 2021-01-01 | 2021-02-08 | 5 |
| Hôpital Saint-François d'Assise, CHU de Québec | Quebec | 2020-12-29 | 2021-04-11 | 9 |
| Hôtel-Dieu de Québec, CHU de Québec | Quebec | 2020-12-29 | 2021-01-05 | <5 |
| IUCPQ: Institut universitaire de cardiologie et de pneumologie de Québec | Quebec | 2020-12-01 | 2020-12-31 | 6 |
| Hôpital du Sacré-Coeur de Montréal | Quebec | 2020-12-22 | 2022-06-24 | 394 |
| Montreal General Hospital | Quebec | 2020-12-05 | 2021-12-30 | 69 |
| Halifax Infirmary | Nova Scotia | 2021-01-26 | 2022-04-24 | 47 |
| Dartmouth General Hospital | Nova Scotia | 2021-04-28 | 2022-04-07 | 22 |
| Cobequid Community Health Centre | Nova Scotia | 2021-01-08 | 2022-04-24 | 11 |

\*The start and end dates correspond to the start and end of the enrolment of COVID-19 tested patients in the **Canadian COVID-19 Emergency Department Rapid Response Network** registry. Eligible patients were those with an index visit date between October 18, 2020, and October 11, 2022. Eligible patients were contacted to obtain consent to participate in this study between November 16, 2021, and August 31, 2023.

**S2 Table.** TRIPOD checklist - Prediction Model Development (in progress)

| Section/Topic | Item | Checklist Item | Page |
| --- | --- | --- | --- |
| **Title and abstract** |  |  |  |
| Title | 1 | Identify the study as developing and/or validating a multivariable prediction model, the target population, and the outcome to be predicted. | 1 |
| Abstract | 2 | Provide a summary of objectives, study design, setting, participants, sample size, predictors, outcome, statistical analysis, results, and conclusions. | 3 |
| **Introduction** |  |  |  |
| Background and objectives | 3a | Explain the medical context (including whether diagnostic or prognostic) and rationale for developing or validating the multivariable prediction model, including references to existing models. | 4-5 |
|  | 3b | Specify the objectives, including whether the study describes the development or validation of the model or both. | 5 |
| **Methods** |  |  |  |
| Source of data | 4a | Describe the study design or source of data (e.g., randomized trial, cohort, or registry data), separately for the development and validation data sets, if applicable. | 6 |
|  | 4b | Specify the key study dates, including start of accrual; end of accrual; and, if applicable, end of follow-up. | 6 |
| Participants | 5a | Specify key elements of the study setting (e.g., primary care, secondary care, general population) including number and location of centres. | 7 |
|  | 5b | Describe eligibility criteria for participants. | 7 |
|  | 5c | Give details of treatments received, if relevant. | 7 |
| Outcome | 6a | Clearly define the outcome that is predicted by the prediction model, including how and when assessed. | 8 |
|  | 6b | Report any actions to blind assessment of the outcome to be predicted. | 8-9 |
| Predictors | 7a | Clearly define all predictors used in developing or validating the multivariable prediction model, including how and when they were measured. | 9 |
|  | 7b | Report any actions to blind assessment of predictors for the outcome and other predictors. | 10 |
| Sample size | 8 | Explain how the study size was arrived at. | 10 |
| Missing data | 9 | Describe how missing data were handled (e.g., complete-case analysis, single imputation, multiple imputation) with details of any imputation method. | 9 |
| Statistical analysis methods | 10a | Describe how predictors were handled in the analyses. | 9 |
|  | 10b | Specify type of model, all model-building procedures (including any predictor selection), and method for internal validation. | 9 |
|  | 10d | Specify all measures used to assess model performance and, if relevant, to compare multiple models. | 9-10 |
| Risk groups | 11 | Provide details on how risk groups were created, if done. | n/a |
| **Results** |  |  |  |
| Participants | 13a | Describe the flow of participants through the study, including the number of participants with and without the outcome and, if applicable, a summary of the follow-up time. A diagram may be helpful. | 11 |
|  | 13b | Describe the characteristics of the participants (basic demographics, clinical features, available predictors), including the number of participants with missing data for predictors and outcome. | 12-13, S4 Table |
| Model development | 14a | Specify the number of participants and outcome events in each analysis. | 13-14 |
|  | 14b | If done, report the unadjusted association between each candidate predictor and outcome. | n/a |
| Model specification | 15a | Present the full prediction model to allow predictions for individuals (i.e., all regression coefficients, and model intercept or baseline survival at a given time point). | n/a |
|  | 15b | Explain how to use the prediction model. | n/a |
| Model performance | 16 | Report performance measures (with CIs) for the prediction model. | 14 |
| **Discussion** |  |  |  |
| Limitations | 18 | Discuss any limitations of the study (such as nonrepresentative sample, few events per predictor, missing data). | 22 |
| Interpretation | 19b | Give an overall interpretation of the results, considering objectives, limitations, and results from similar studies, and other relevant evidence. | 15-16, 22-23 |
| Implications | 20 | Discuss the potential clinical use of the model and implications for future research. | 23 |
| **Other information** |  |  |  |
| Supplementary information | 21 | Provide information about the availability of supplementary resources, such as study protocol, Web calculator, and data sets. | 40 |
| Funding | 22 | Financial Statement disclosure | 39 |

**S3 Table.** GRIPP2-Short Form Checklist for the Reporting of Patient Engagement in Research.

| Section and topic | Item | Reported on page No |
| --- | --- | --- |
| 1: Aim<br>Report the aim of patient and public involvement in the study | To collaboratively involve patients as research partners at all stages in the development of this project. *“Fifth, this study was developed with the participation of patient partners who provided guidance in its development, its conduct and interpretation.”* | 16 |
| 2: Methods<br>Provide a clear description of the methods used for patient and public involvement in the study | CCEDRRN's Patient Engagement Committee consisted of 11 members from 6 provinces across Canada (BC, AB, SK, MN, ON, NB). Members have lived experience with COVID-19 and PCC and were engaged from both CCEDRRN's and this study's inception onwards. They participated in generating the original research question, grant writing, edited consent and data collection forms to ensure readability and acceptability in French and English and across diverse social, cultural, and ethnic contexts, assisted with developing and refining study questions, interpreted study findings, and reviewed and edited the manuscript. *“We developed the PCCAQ based on the WHO PCC case definition and case report form in collaboration with patient partners, PCC experts, emergency physicians, rehabilitation specialists, and public health policy makers. We piloted a first version of the PCCAQ (PCCAQ version 1.0, S1a File) in English and French with CCEDRRN patient partners and the first 100 participants. Then, we used this version of the PCCAQ with all patients whose index visit occurred between October 18, 2020, and February 28, 2022. For all patients with index visits after February 28, 2022, we used a new and simplified version of the PCCAQ that was improved to help patients determine the duration of their symptoms using open-ended questions instead of asking them to categorize the duration of their symptoms using predetermined multiple-choice options (PCCAQ version 2.0, S1b File)”.* | 8 |
| 3: Study results<br>Outcomes—Report the results of patient and public involvement in the study, including both positive and negative outcomes | Among the 11 original patient partners, four patient partners were more active in this project.<br>Positive outcomes: (1) generation of the original patient-oriented study question; (2) timely grant writing for a patient-oriented research project; (3) consent form and data collection improvements and adaptations to patient's needs; (4) networking and knowledge dissemination; (5) raised awareness about the experience and needs of Long COVID (PCC) patients; (6) raised awareness about Long COVID (PCC) among team members and clinicians and its perception in the general public. (7) Awareness about the sensitivities in how science is reported and its impact on patients' wellbeing.<br>Negative outcomes: (1) The pandemic negatively impacted patient partners' and researchers' experience in | S3 Table |
|  | leading patient-oriented research. Leading patient-oriented research through a pandemic was not easy. Even though virtual meetings facilitated people getting together, it also added stress on patient partners and researchers. (2) The speed at which evidence generation was needed during the pandemic added stress on patients and researchers. The need to generate timely evidence clashed with the time-investment needed to build trusting and meaningful relationships with patient partners. (3) Patient partners representing marginalized groups and populations were even harder to recruit and include during the pandemic. (4) Finding the delicate balance between patient engagement and patient advocacy was a challenge. (5) Finding the delicate balance in interpreting data in the most objective and neutral way was also a challenge. (6) Ethics committees in all provinces (except BC) did not allow consent to be obtained from patients who could not express themselves in English or French. This forced our team to exclude patients who could not read or speak English or French in all provinces (excluding BC where proxy consent and data collection was allowed). |  |
| 4: Discussion and conclusions<br>Outcomes—Comment on the extent to which patient and public involvement influenced the study overall. Describe positive and negative effects | Positive effects: (1) More relevant research question; (2) Funding was obtained through the Canadian Institutes for Health Research with the participation of four patient partners; (3) Better data collection tools; (4) Balanced interpretation of the data; (5) New lasting collaborations and relationships; (6) Better knowledge translation and dissemination; (7) New opportunities for future research; (8) Increased awareness and empathy for patients living with a new illness.<br>Negative effects: (1) Time consuming; (2) Increased cost and need for additional funding; (3) Moral fatigue and weariness; (4) Added stress and burden on the research process and difficulty in the sustainability of the patient-engagement process without adequate research funding; (5) Grief and mourning due to the loss of a patient partner to illness. | S3 Table |
| 5: Reflections<br>*Comment critically on the study, reflecting on the things that went well and those that did not, so others can learn from this experience* | This patient-oriented research project was a great experience. It was a great example of the benefits of bringing patients and researchers together as the question for this project emerged from CCEDRRN's patient engagement committee. The funding for this project was obtained with the participation of patient partners. The conduct of the study was improved with patient partners' participation. In the future, we need more funding to continue the ongoing activities and relationship-building needed to plan future studies. Less administrative barriers are needed for more timely involvement of patient partners in the future. Additionally, better involvement of marginalized populations as patient partners will take more time and will need more preparation and training. | S3 Table |

**S1a File. Post COVID-19 Condition Assessment Questionnaire (PCCAQ) Version 1.0\***

*Note: This was the questionnaire used with patients with index visits from October 18, 2021, to February 28, 2022.

1. **Did you visit the emergency department since you went on YYYY/MM/DD?**

No

Yes

Prefer not to answer

If yes, a. How many times did you go back to the emergency department? *(number)* b. For your first revisit to the ED, do you remember how long it was after your initial visit of the YYYY/MM/DD? A couple of days Within 1 week Within 2 weeks Within 1 month Between 2 to 6 months More than 6 months c. Were you admitted to the hospital during that visit? Yes No Prefer not to answer

3. Since the acute health problem that brought you to the emergency department on YYYY/MM/DD, have you been more short of breath?

No, not at all

Yes, and it is still present

Yes, sometimes and it is still present

Yes, but not anymore

I don’t know

Prefer not to answer

If yes, and it is still present / yes, sometimes and it is still present / yes, but not anymore

a. When do / did you experience shortness of breath? At rest During low to moderate intensity activity (ex. walking, cleaning, doing the dishes) During high intensity activity (ex. exercising, jogging, biking)

If yes, and it is still present / yes, sometimes and it is still present / yes, but not anymore

b. When did the shortness of breath start? (calendar) If yes, but not anymore

i. How long did the shortness of breath last? A couple of days 1 week 2 weeks Between 1 and 2 months Between 2 to 6 months More than 6 months

4. Since the acute health problem that brought you to the emergency department on YYYY/MM/DD, have you had any new and persisting pain?

No, not at all

Yes, and it is still present

Yes, sometimes and it is still present

Yes, but not anymore

I don’t know

Prefer not to answer

If yes, and it is still present / yes, sometimes and it is still present / yes, but not anymore

a. Where is / was the main source of your pain?

- Abdominal pain
- Chest pain
- Back pain
- Sore throat
- Stomach ache
- Headache or Migraine
- Joints pain
- Pain while breathing
- Pain when coughing
- Muscle pain
- Pain in the legs
- Pain in the arms
- Pain when swallowing
- Generalized pain (non-specified)
- Other
b. When did the pain start? *(calendar)* If yes, but not anymore

i. How long did the pain last? A couple of days 1 week 2 weeks Between 1 and 2 months Between 2 to 6 months More than 6 months

5. Since the acute health problem that brought you to the emergency department on YYYY/MM/DD, have you had a new and persistent cough?

No, not at all

Yes, and it is still present

Yes, sometimes and it is still present

Yes, but not anymore

I don’t know

Prefer not to answer

If yes, and it is still present / yes, sometimes and it is still present / yes, but not anymore

a. When did the cough start? *(calendar)* If yes, but not anymore

i. How long did the cough last? A couple of days 1 week 2 weeks Between 1 and 2 months Between 2 to 6 months More than 6 months

6. Since the acute health problem that brought you to the emergency department on YYYY/MM/DD, have you had a decreased sense of smell?

No, not at all

Yes, and it is still present

Yes, sometimes and it is still present

Yes, but not anymore

I don’t know

Prefer not to answer

If yes, and it is still present / yes, sometimes and it is still present / yes, but not anymore

a. When did your decreased sense of smell start? *(calendar)* If yes, but not anymore

i. How long did your decreased sense of smell last? A couple of days 1 week 2 weeks Between 1 and 2 months Between 2 to 6 months More than 6 months

7. Since the acute health problem that brought you to the emergency department on YYYY/MM/DD, have you had a decreased sense of taste?

No, not at all

Yes, and it is still present

Yes, sometimes and it is still present

Yes, but not anymore

I don’t know

Prefer not to answer

If yes, and it is still present / yes, sometimes and it is still present / yes, but not anymore

a. When did your decreased sense of taste start? *(calendar)* If yes, but not anymore

i. How long did your decreased sense of taste last? A couple of days 1 week 2 weeks Between 1 and 2 months Between 2 to 6 months More than 6 months

8. Since the acute health problem that brought you to the emergency department on YYYY/MM/DD, has your sleep been different?

No, not at all

Yes, and it is still present

Yes, sometimes and it is still present

Yes, but not anymore

I don’t know

Prefer not to answer

If yes, and it is still present / yes, sometimes and it is still present / yes, but not anymore

a. When did your sleep change? *(calendar)* If yes, but not anymore

i. For how long was your sleep different? A couple of days 1 week 2 weeks Between 1 and 2 months Between 2 to 6 months More than 6 months

9. Since the acute health problem that brought you to the emergency department on YYYY/MM/DD, have you felt dizzy or lightheaded?

No, not at all

Yes, and it is still present

Yes, sometimes and it is still present

Yes, but not anymore

I don’t know

Prefer not to answer

If yes, and it is still present / yes, sometimes and it is still present / yes, but not anymore

a. When did your dizziness or lightheadedness start? *(calendar)* If yes, but not anymore

i. How long did your dizziness or lightheadedness last? A couple of days 1 week 2 weeks Between 1 and 2 months Between 2 to 6 months More than 6 months

10. Since the acute health problem that brought you to the emergency department on YYYY/MM/DD, has it been harder to concentrate?

No, not at all

Yes, and it is still present

Yes, sometimes and it is still present

Yes, but not anymore

I don’t know

Prefer not to answer

If yes, and it is still present / yes, sometimes and it is still present / yes, but not anymore

a. When did it start to get harder to concentrate? *(calendar)* If yes, but not anymore

i. For how long was it harder to concentrate? A couple of days 1 week 2 weeks Between 1 and 2 months Between 2 to 6 months More than 6 months

11. Since the acute health problem that brought you to the emergency department on YYYY/MM/DD, has it been harder to remember things?

No, not at all

Yes, and it is still present

Yes, sometimes and it is still present

Yes, but not anymore

I don’t know

Prefer not to answer

If yes, and it is still present / yes, sometimes and it is still present / yes, but not anymore

a. When did it start to get harder to remember things? *(calendar)* If yes, but not anymore

i. For how long was it harder to remember things? A couple of days 1 week 2 weeks Between 1 and 2 months Between 2 to 6 months More than 6 months

12. Since the acute health problem that brought you to the emergency department on YYYY/MM/DD, have you felt unusually tired after a physical, a mental or an emotional effort?

No, not at all

Yes, and it is still present

Yes, sometimes and it is still present

Yes, but not anymore

I don’t know

Prefer not to answer

If yes, and it is still present / yes, sometimes and it is still present / yes, but not anymore

a. When did this unusual tiredness start? *(calendar)* If yes, but not anymore

i. How long did this unusual tiredness last? A couple of days 1 week 2 weeks Between 1 and 2 months Between 2 to 6 months More than 6 months

13. Since the acute health problem that brought you to the emergency department on YYYY/MM/DD, have you had other symptoms than the ones we discussed that you would like to share today?

No Yes

Prefer not to answer

If yes,

a. Which one affects / affected you the most?

- Anxiety
- Behaviour change
- Can’t move and/or feel one side of body or face
- Constipation
- Depressed mood
- Diarrhea
- Dry Mouth
- Dysmenorrhea
- Eye dryness
- Fainting/blackouts
- Fever
- Food sensitivities
- Hot flashes
- Jerking of limbs
- Joint swelling
- Loss of appetite
- Loss of interest/pleasure
- Loss of teeth
- Lumpy lesions: (purple/pink/bluish) on toes/COVID toes
- Nausea/vomiting
- Night sweats
- Numbness or tingling
- Persistent fatigue
- Problems hearing
- Problems passing urine
- Problems seeing
- Problems swallowing
- Problems with balance
- Problems with gait/falls
- Ringing in ears
- Seizures
- Skin rash
- Slowness of movement
- Stiffness of muscles
- Tremors
- Weakness in limbs
- Weight loss
- Erectile dysfunction
- Hallucinations

If yes,

b. When did this symptom start? *(calendar)*

i. How long did this symptom last? A couple of days 1 week 2 weeks Between 1 and 2 months Between 2 to 6 months More than 6 months Still present

**S1b File. Post COVID-19 Condition Assessment Questionnaire (PCCAQ) Version 2.0\***

*Note: This was the questionnaire used with patients with index visit between March 1, 2022 and October 11, 2022.

**1. Did you visit the emergency department since you went on YYYY/MM/DD?**

☐No
☐Yes
☐Prefer not to answer

If yes,

**How many times did you go back to the emergency department? *(number)***

**2. Since the acute health problem that brought you to the emergency department on YYYY/MM/DD, have you been more short of breath?**

☐No, not at all
☐Yes, and it is still present
☐Yes, sometimes and it is still present
☐Yes, but not anymore
☐I don’t know

☐Prefer not to answer

If yes, and it is still present / yes, sometimes and it is still present / yes, but not anymore

**When do / did you experience shortness of breath?**

☐ At rest
☐ During low to moderate intensity activity (ex. walking, cleaning, doing the dishes)
☐ During high intensity activity (ex. exercising, jogging, biking)

If yes, and it is still present / yes, sometimes and it is still present / yes, but not anymore

**When did the shortness of breath start? *(calendar)***

If yes, but not anymore

**Did your symptom last 2 months or longer?**

☐ Yes
☐ No

If yes,

**How many months did your symptom last?**

**3. Since the acute health problem that brought you to the emergency department on YYYY/MM/DD, have you had any new and persisting pain?**

☐No, not at all
☐Yes, and it is still present
☐Yes, sometimes and it is still present
☐Yes, but not anymore
☐I don’t know
☐Prefer not to answer

If yes, and it is still present / yes, sometimes and it is still present / yes, but not anymore

**3a. Where is / was the main source of your pain?**

- Abdominal pain
- Chest pain
- Back pain
- Sore throat
- Stomach ache
- Headache or Migraine
- Joints pain
- Pain while breathing
- Pain when coughing
- Muscle pain
- Pain in the legs
- Pain in the arms
- Pain when swallowing
- Generalized pain (non-specified)
- Other

**3b**. **When did the pain start?** *(calendar)*

If yes, but not anymore

**Did your symptom last 2 months or longer?**

☐ Yes
☐ No

If yes,

**How many months did your symptom last?**

**4. Since the acute health problem that brought you to the emergency department on YYYY/MM/DD, have you had a new and persistent cough?**

☐No, not at all
☐Yes, and it is still present
☐Yes, sometimes and it is still present
☐Yes, but not anymore
☐I don’t know
☐Prefer not to answer

If yes, and it is still present / yes, sometimes and it is still present / yes, but not anymore

**When did the cough start?** *(calendar)*

If yes, but not anymore

**Did your symptom last 2 months or longer?**

☐ Yes
☐ No

If yes,

**How many months did your symptom last?**

**5. Since the acute health problem that brought you to the emergency department on YYYY/MM/DD, have you had a decreased sense of smell?**

☐No, not at all
☐Yes, and it is still present
☐Yes, sometimes and it is still present
☐Yes, but not anymore
☐I don’t know
☐Prefer not to answer

If yes, and it is still present / yes, sometimes and it is still present / yes, but not anymore

**When did your decreased sense of smell start?** *(calendar)*

If yes, but not anymore

**Did your symptom last 2 months or longer?**

☐ Yes
☐ No

If yes,

**How many months did your symptom last?**

**6. Since the acute health problem that brought you to the emergency department on YYYY/MM/DD, have you had a decreased sense of taste?**

☐No, not at all
☐Yes, and it is still present
☐Yes, sometimes and it is still present
☐Yes, but not anymore
☐I don’t know
☐Prefer not to answer

If yes, and it is still present / yes, sometimes and it is still present / yes, but not anymore

**When did your decreased sense of taste start?** *(calendar)*

If yes, but not anymore

**Did your symptom last 2 months or longer?**

☐ Yes
☐ No

If yes,

**How many months did your symptom last?**

**7. Since the acute health problem that brought you to the emergency department on YYYY/MM/DD, has your sleep been different?**

☐No, not at all
☐Yes, and it is still present
☐Yes, sometimes and it is still present
☐Yes, but not anymore
☐I don’t know
☐Prefer not to answer

If yes, and it is still present / yes, sometimes and it is still present / yes, but not anymore

**When did your sleep change?** *(calendar)*

If yes, but not anymore

**Did your symptom last 2 months or longer?**

☐ Yes
☐ No

If yes,

**How many months did your symptom last?**

**8. Since the acute health problem that brought you to the emergency department on YYYY/MM/DD, have you felt dizzy or lightheaded?**

☐No, not at all
☐Yes, and it is still present
☐Yes, sometimes and it is still present
☐Yes, but not anymore
☐I don’t know
☐Prefer not to answer

If yes, and it is still present / yes, sometimes and it is still present / yes, but not anymore

**When did your dizziness or lightheadedness start?** *(calendar)*

If yes, but not anymore

**Did your symptom last 2 months or longer?**

☐ Yes
☐ No

If yes,

**How many months did your symptom last?**

**9. Since the acute health problem that brought you to the emergency department on YYYY/MM/DD, has it been harder to concentrate?**

☐No, not at all
☐Yes, and it is still present
☐Yes, sometimes and it is still present
☐Yes, but not anymore
☐I don’t know
☐Prefer not to answer

If yes, and it is still present / yes, sometimes and it is still present / yes, but not anymore

**When did it start to get harder to concentrate?** *(calendar)*

If yes, but not anymore

**Did your symptom last 2 months or longer?**

☐ Yes
☐ No

If yes,

**How many months did your symptom last?**

☐More than 6 months

**10. Since the acute health problem that brought you to the emergency department on YYYY/MM/DD, has it been harder to remember things?**

☐No, not at all
☐Yes, and it is still present
☐Yes, sometimes and it is still present
☐Yes, but not anymore
☐I don’t know
☐Prefer not to answer

If yes, and it is still present / yes, sometimes and it is still present / yes, but not anymore

**When did it start to get harder to remember things?** *(calendar)*

If yes, but not anymore

**Did your symptom last 2 months or longer?**

☐ Yes
☐ No

If yes,

**How many months did your symptom last?**

**11. Since the acute health problem that brought you to the emergency department on YYYY/MM/DD, have you felt unusually tired after a physical, a mental or an emotional effort?**

☐No, not at all
☐Yes, and it is still present
☐Yes, sometimes and it is still present
☐Yes, but not anymore
☐I don’t know
☐Prefer not to answer

If yes, and it is still present / yes, sometimes and it is still present / yes, but not anymore

**When did this unusual tiredness start?** *(calendar)*

If yes, but not anymore

**Did your symptom last 2 months or longer?**

☐ Yes
☐ No

If yes,

**How many months did your symptom last?**

**12. Since the acute health problem that brought you to the emergency department on YYYY/MM/DD, have you had other symptoms than the ones we discussed that you would like to share today?**

☐No
☐Yes
☐Prefer not to answer

If yes,

**Which one affects / affected you the most**?

- Anxiety
- Behaviour change
- Can’t move and/or feel one side of body or face
- Constipation
- Depressed mood
- Diarrhea
- Dry Mouth
- Dysmenorrhea
- Eye dryness
- Fainting/blackouts
- Fever
- Food sensitivities
- Hot flashes
- Jerking of limbs
- Joint swelling
- Loss of appetite
- Loss of interest/pleasure
- Loss of teeth
- Lumpy lesions: (purple/pink/bluish) on toes/COVID toes
- Nausea/vomiting
- Night sweats
- Numbness or tingling
- Persistent fatigue
- Problems hearing
- Problems passing urine
- Problems seeing
- Problems swallowing
- Problems with balance
- Problems with gait/falls
- Ringing in ears
- Seizures
- Skin rash
- Slowness of movement
- Stiffness of muscles
- Tremors
- Weakness in limbs
- Weight loss
- Erectile dysfunction
- Hallucinations

If yes,

**When did this symptom start?** *(calendar)*

**Did your symptom last 2 months or longer?**

☐ Yes
☐ No

If yes,

**How many months did your symptom last?**

**S4 Table.** Candidate variables for the regression model.

| Variable | Definition | N (%) Missing |
| --- | --- | --- |
| Age | Age in years | 0 (0) |
| Sex | Male, Female | 0 (0) |
| Pandemic wave | Pre-Omicron (before November 28, 2021), Omicron (On or after November 28, 2021) | 0 (0) |
| Vaccination status | Patient has received any type of vaccination 14 days prior to their index ED visit. 0/1 dose vs 2 or more doses | 0 (0) |
| **Emergency department variables** |  |  |
| ED arrival mode |  | 0 (0) |
| Ambulance | arrived by ambulance |  |
| Self/police | self-transported or transported to ED by police |  |
| Arrival heart rate | beats/minute | 139 (2.3) |
| Arrival respiratory rate | breaths/minute | 94 (1.5) |
| Arrival systolic blood pressure | mmHg | 139 (2.3) |
| Arrival temperature | Celsius | 194 (3.2) |
| CTAS | Canadian Triage and Acuity Scale | 14 (0.2) |
| Respiratory distress | Increased work of breathing documented by treating clinician and Patient-reported symptom of shortness of breath as documented by treating clinician | 0 (0) |
| Supplemental oxygen delivered in the ED | Yes/No | 0 (0) |
| Mode of oxygen delivery combined with oxygen received in the ED | Documented mode of oxygen delivery, combined with maximum oxygen received if given nasal prongs. Most invasive method of oxygen delivery selected if more than one documented. | 0 (0) |
|  | Intubation<br>BiPap + CPAP + HFNO<br>Facemask + simple rebreather + non-rebreather + nasal prongs with > 6L/min<br>Nasal prongs <5L/min<br>No oxygen |  |
| Current tobacco user | Documented current tobacco use | 0 (0) |
| Current illicit user | Documented methamphetamine, opioid or other illicit drug use | 0 (0) |
| Current Alcohol misuse | Documented current alcohol misuse | 0 (0) |
| Admitted | Yes/No | 0 (0) |
| COVID symptoms |  |  |
| Abdominal pain | Patient-reported symptom as documented by treating clinician | 0 (0) |
| Altered consciousness/confusion | Patient-reported symptom as documented by treating clinician | 0 (0) |
| Chest pain (Includes discomfort or tightness) | Patient-reported symptom as documented by treating clinician | 0 (0) |
| Chills | Patient-reported symptom as documented by treating clinician | 0 (0) |
| Cough | Patient-reported symptom as documented by treating clinician | 0 (0) |
| Diarrhea | Patient-reported symptom as documented by treating clinician | 0 (0) |
| Dizziness/Vertigo | Patient-reported symptom as documented by treating clinician | 0 (0) |
| Dysgeusia/anosmia | Patient-reported symptom as documented by treating clinician | 0 (0) |
| Fatigue/malaise | Patient-reported symptom as documented by treating clinician | 0 (0) |
| Fever | Patient-reported symptom as documented by treating clinician | 0 (0) |
| Headache | Patient-reported symptom as documented by treating clinician | 0 (0) |
| Muscle aches (myalgia) | Patient-reported symptom as documented by treating clinician | 0 (0) |
| Nausea/vomiting | Patient-reported symptom as documented by treating clinician | 0 (0) |
| Runny nose (rhinorrhea) | Patient-reported symptom as documented by treating clinician | 0 (0) |
| Seizures | Patient-reported symptom as documented by treating clinician | 0 (0) |
| Shortness of breath (dyspnea) | Patient-reported symptom as documented by treating clinician | 0 (0) |
| Skin rash | Patient-reported symptom as documented by treating clinician | 0 (0) |
| Skin ulcers | Patient-reported symptom as documented by treating clinician | 0 (0) |
| Comorbid Conditions |  |  |
| Active malignant neoplasm (cancer) | The diagnosis, conditions, problem or circumstances for the patient's ED visit that is in addition to the main diagnosis. | 0 (0) |
| Asthma | The diagnosis, conditions, problem or circumstances for the patient's ED visit that is in addition to the main diagnosis. | 0 (0) |
| Atrial Fibrillation | The diagnosis, conditions, problem or circumstances for the patient's ED visit that is in addition to the main diagnosis. | 0 (0) |
| Chronic kidney disease/Dialysis | The diagnosis, conditions, problem or circumstances for the patient's ED visit that is in addition to the main diagnosis. | 0 (0) |
| Chronic lung disease/Pulmonary fibrosis (not asthma, e.g., COPD, IPF) | The diagnosis, conditions, problem or circumstances for the patient's ED visit that is in addition to the main diagnosis. | 0 (0) |
| Chronic neuro disorder (not dementia, e.g., stroke/TIA, seizure disorder) | The diagnosis, conditions, problem or circumstances for the patient's ED visit that is in addition to the main diagnosis. | 0 (0) |
| Congestive heart failure | The diagnosis, conditions, problem or circumstances for the patient's ED visit that is in addition to the main diagnosis | 0 (0) |
| Coronary artery disease | The diagnosis, conditions, problem or circumstances for the patient's ED visit that is in addition to the main diagnosis | 0 (0) |
| Diabetes | The diagnosis, conditions, problem or circumstances for the patient's ED visit that is in addition to the main diagnosis | 0 (0) |
| Dyslipidemia | The diagnosis, conditions, problem or circumstances for the patient's ED visit that is in addition to the main diagnosis | 0 (0) |
| Hypertension | The diagnosis, conditions, problem or circumstances for the patient's ED visit that is in addition to the main diagnosis | 0 (0) |
| Hypothyroidism | The diagnosis, conditions, problem or circumstances for the patient's ED visit that is in addition to the main diagnosis | 0 (0) |
| Mild/moderate/severe liver disease | The diagnosis, conditions, problem or circumstances for the patient's ED visit that is in addition to the main diagnosis | 0 (0) |
| Obesity (clinical impression) | The diagnosis, conditions, problem or circumstances for the patient's ED visit that is in addition to the main diagnosis | 0 (0) |
| Organ transplant | The diagnosis, conditions, problem or circumstances for the patient's ED visit that is in addition to the main diagnosis | 0 (0) |
| Past malignant neoplasm (cancer) | The diagnosis, conditions, problem or circumstances for the patient's ED visit that is in addition to the main diagnosis | 0 (0) |
| Psychiatric Condition/Mental Health Diagnosis | The diagnosis, conditions, problem or circumstances for the patient's ED visit that is in addition to the main diagnosis | 0 (0) |
| Rheumatologic disorder | The diagnosis, conditions, problem or circumstances for the patient's ED visit that is in addition to the main diagnosis | 0 (0) |
| **Self-reported socioeconomic variables collected during telephone survey** |  |  |
| Race | The race patient most identified with | 63 (1.0) |
|  | Arab/Middle Eastern<br>Black<br>East/Southeast Asian<br>Indigenous<br>Latin American<br>South Asian<br>White<br>Other<br>Unknown |  |
| Education | Patient's highest level of education completed | 190 (3.1) |
|  | Below high school diploma<br>High school diploma<br>Trade certification or diploma<br>University certificate or diploma<br>University bachelor level or above<br>Unknown |  |

**S5 Table.** Internal bootstrap validation result of the final model with 1000 bootstrap samples.

| **Variable** | **Times retained** | **Included in the reduced model** |
| --- | --- | --- |
| Sex | 1000 | Yes |
| South Asian | 930 | Yes |
| Arrival RR (breaths/min) | 888 | Yes |
| COVID-19 symptoms: Sputum production | 810 | Yes |
| Age | 746 | No |
| COVID-19 symptoms: Fatigue/ malaise | 717 | Yes |
| Comorbid condition: Rheumatologic disorder | 685 | Yes |
| COVID-19 symptoms: Dizziness/ Vertigo | 655 | Yes |
| COVID-19 symptoms: Diarrhea | 630 | Yes |
| COVID-19 symptoms: Chest pain | 591 | Yes |
| Comorbid condition: Psychiatric Condition/Mental Health Diagnosis | 578 | Yes |
| Education | 560 | No |

**S1 Fig.**
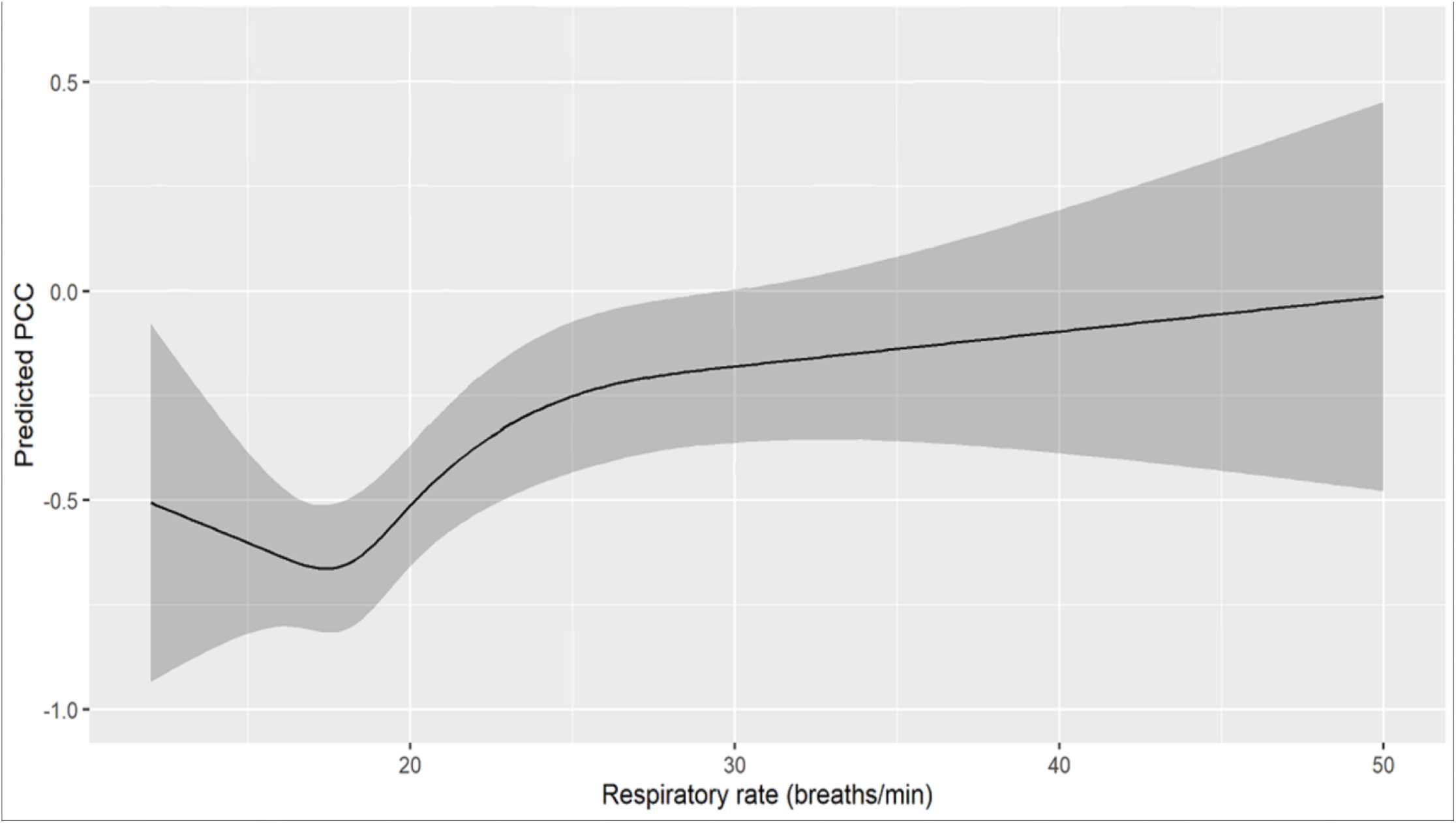
Predicted log odds of Post Covid-19 Condition (PCC) over splined respiratory rate in the final model.

**S2 File: Contributors to the Canadian COVID-19 Emergency Department Rapid Response Network**

1. Purpose

This supplementary table provides details of the support staff at each of the participating institutions in the Canadian COVID-19 Emergency Department Rapid Response Network. This supplementary document should be attached to each peer-reviewed manuscript after the methods manuscript (M1). The purpose is to ensure research staffs and lead coordinators are appropriately recognized for their contributions to the network.

2. List of Support Staff

**Table 1.**
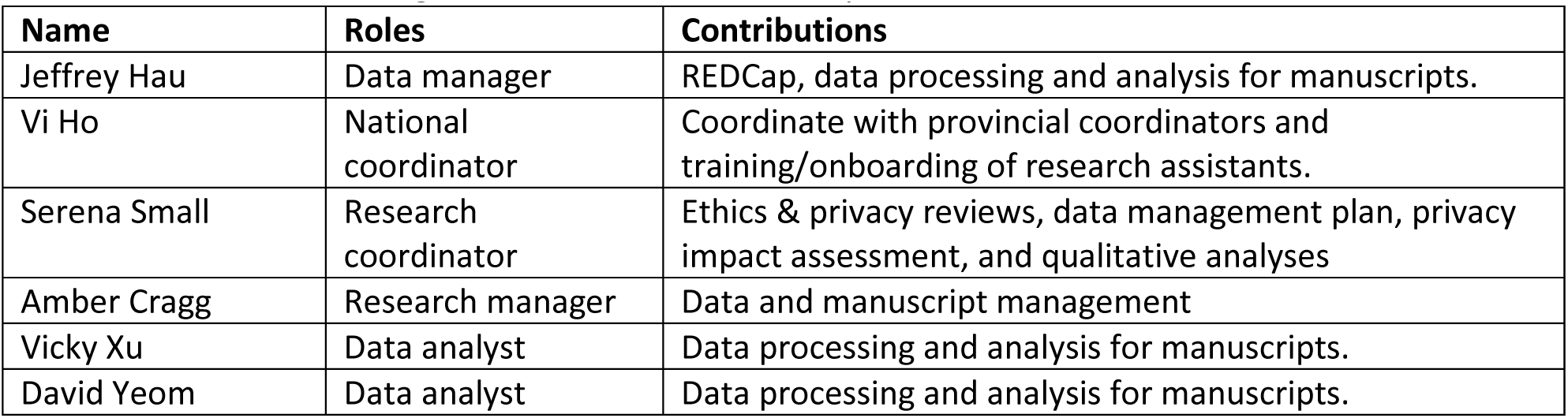
Network coordinating center staff at the University of British Columbia.

**Table 2.**
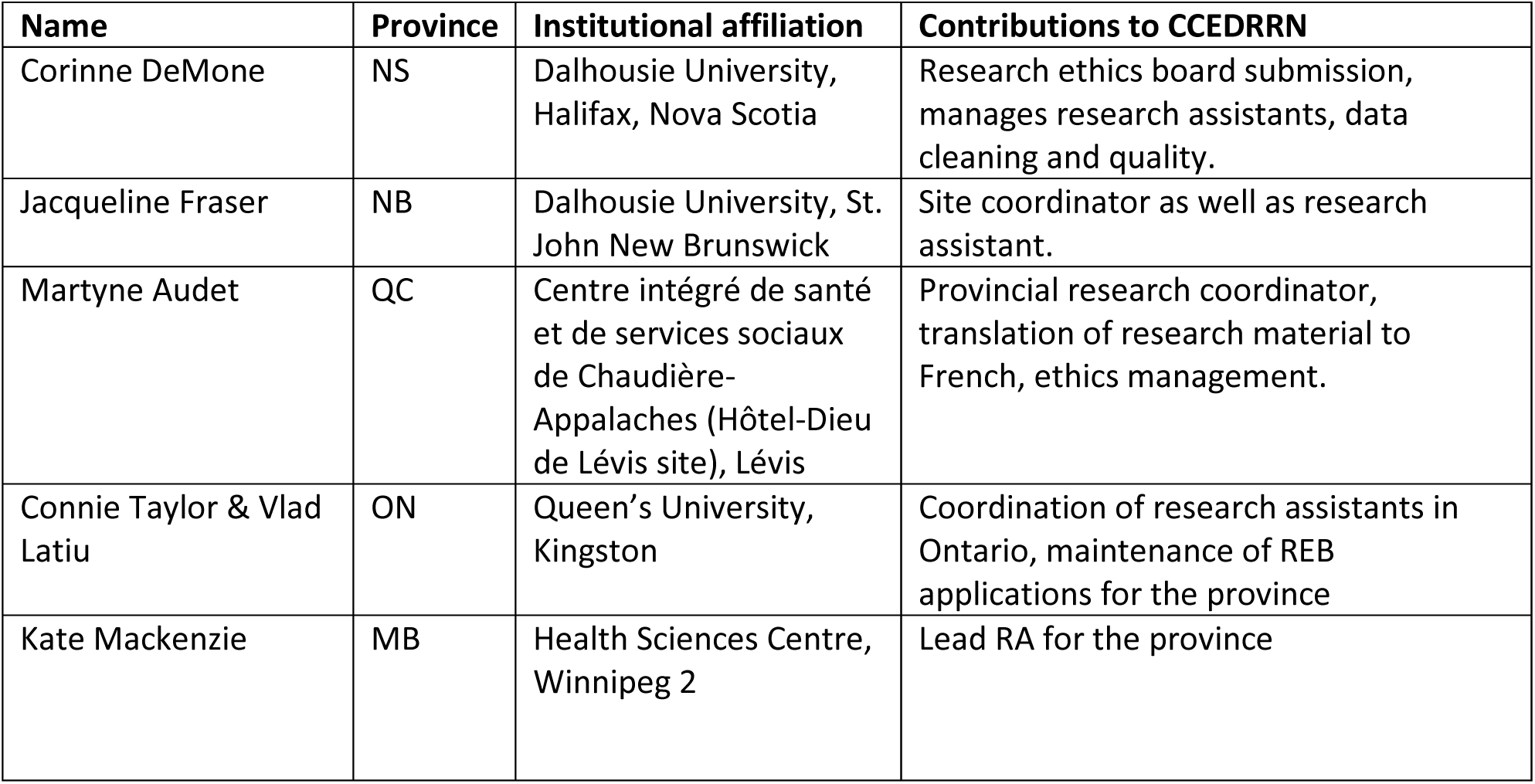
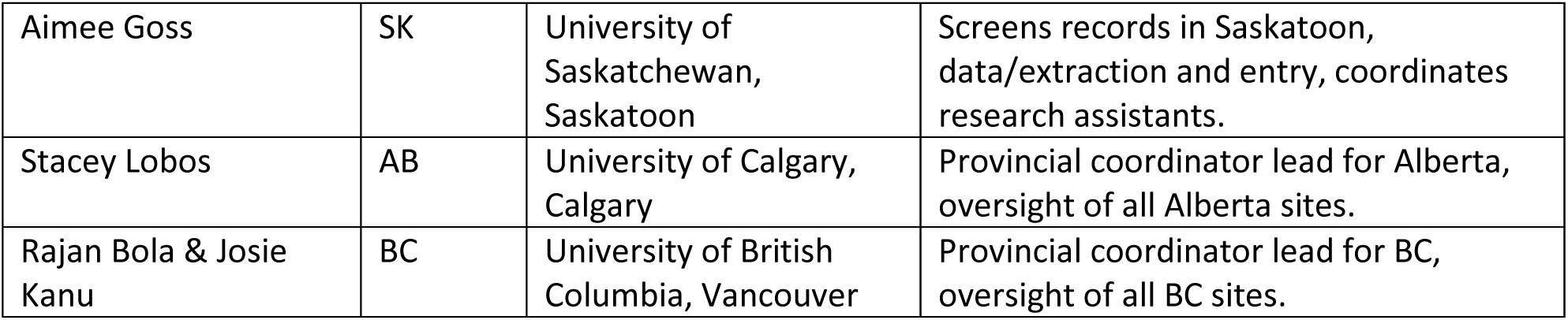
Provincial Coordinators.

**Table 3.**
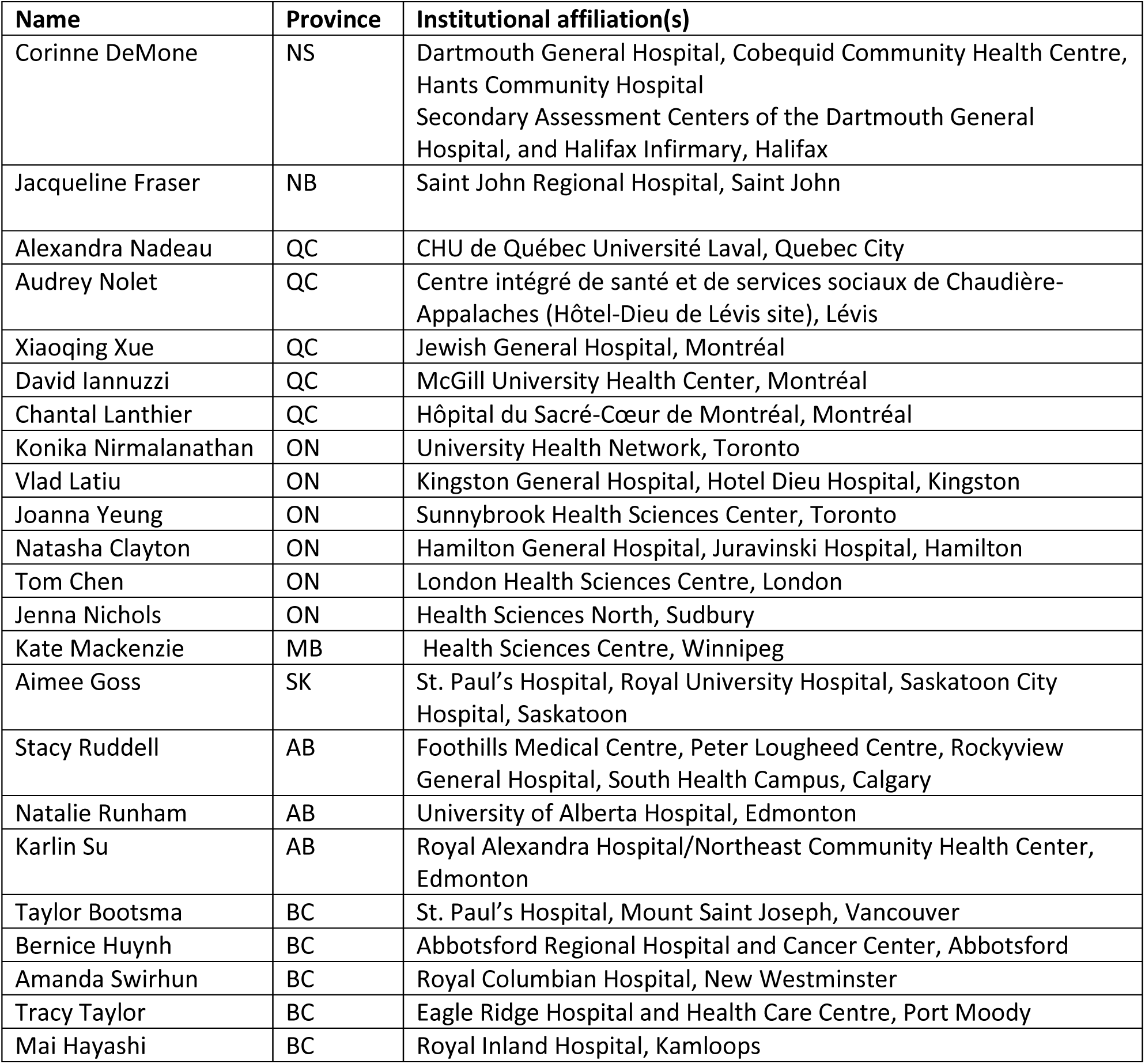
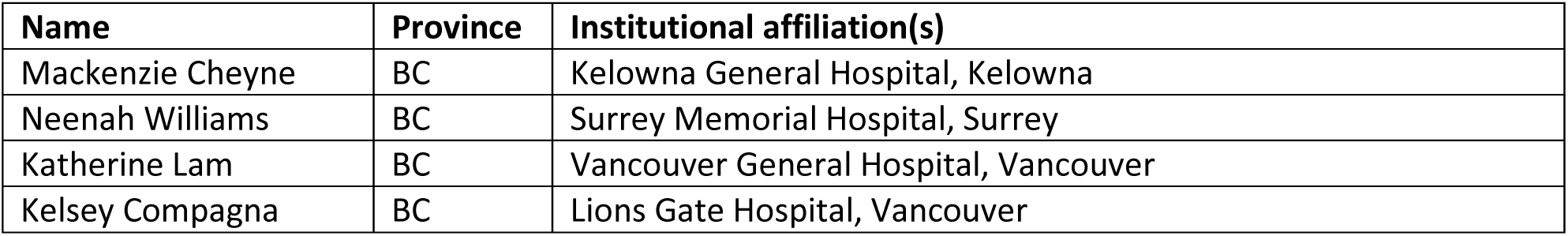
Institutional research assistant (RA) leads Institutional RA leads are responsible for data extraction and integrity, communication with provincial leads.

**Table 4.**
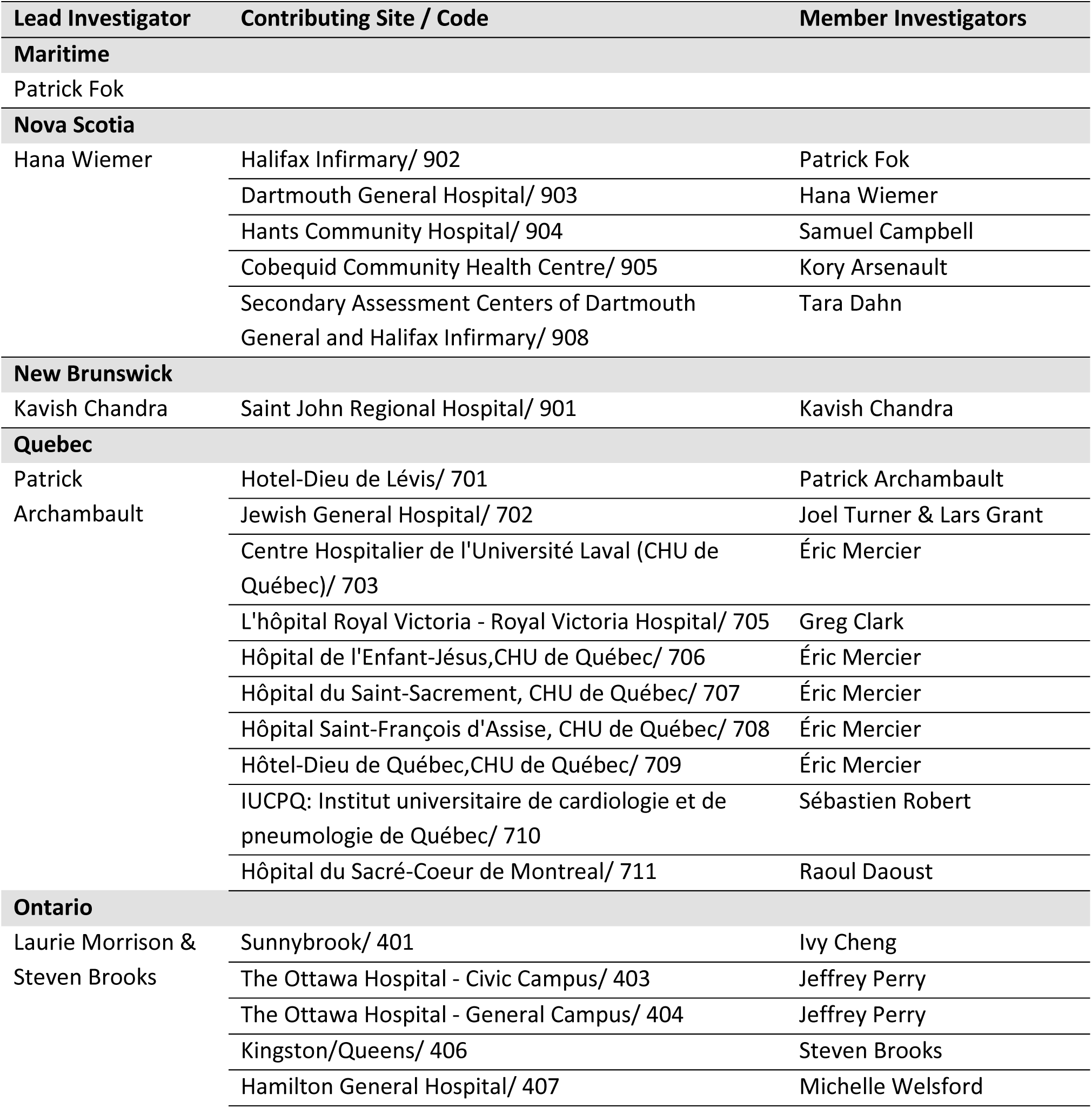
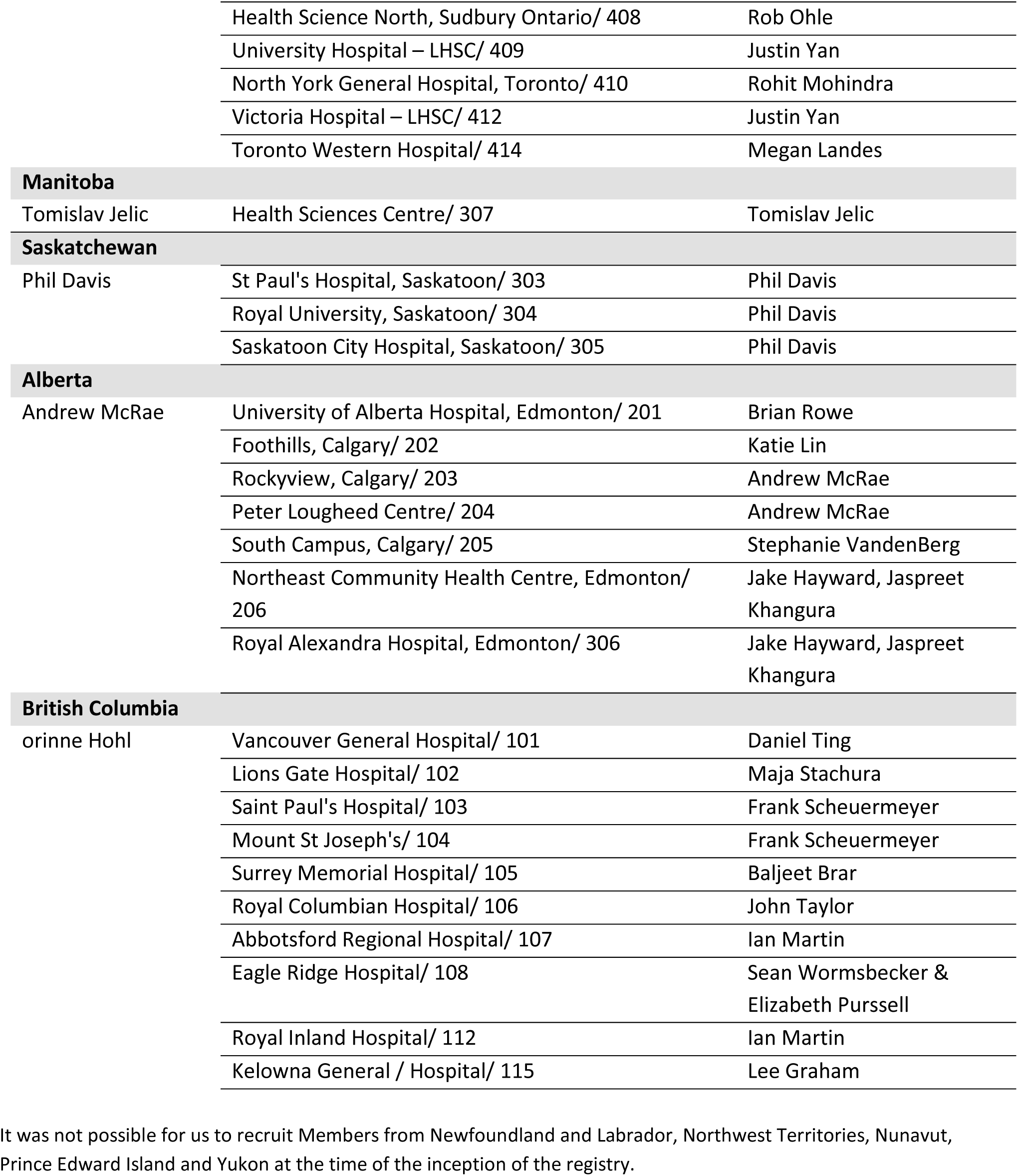
Contributing Study Sites and Investigators.

